# Does the method matter? Evaluating the effectiveness, efficiency and ease of hearing-aid gain self-adjustment

**DOI:** 10.64898/2026.06.11.26355463

**Authors:** Janin Benecke, William M. Whitmer

## Abstract

In conventional hearing-aid personalisation, clinicians cannot hear what their patients hear, and patients cannot often reliably detect or describe what they hear. Self-adjustment can reduce this issue but requires user controls that adjust hearing-aid signal processing parameters to be effective, efficient and easy. In this study, we explored (a) the roles of interface complexity and stimulus type in the self-adjustment of linear frequency-dependent gain, and (b) how well individuals can adjust one sound to match another to assess the same interfaces and stimuli. Adult hearing-aid users with mild to moderate symmetrical sensorineural hearing loss repeatedly adjusted the gain (a) to their preference from individual prescription (*n* = 41) and (b) to match their previous preferences from a random starting point (*n* = 32) using three interfaces representing different bass/mid/treble configurations and three stimuli (music, speech and speech-in-noise). Exploratory clustering of the interindividual preferred-gain variability suggested three patterns of deviation from initial prescription: increased relative bass, overall gain reduction, and close to initial prescription. There were no substantial effects of interface nor stimulus on self-adjustment reliability (median σ = 2.8 dB), whereas absolute sound-matching error increased with increasing interface complexity and centre frequency. Neither individual matching accuracy nor questionnaire responses predicted either self-adjusted gains or reliability. Overall, these results show that many – but not all – hearing-aid users can adjust gains with reasonable reliability, and while it can be difficult to predict the behaviour from the individual, the individual applies a similar self-adjustment behaviour across different interfaces and stimuli.

## INTRODUCTION

Frequency-dependent gain is a fundamental hearing-aid parameter. Amplification provides audibility, and thereby shapes how a device sounds, which in turn may affect a user’s satisfaction with and use of their hearing aids (McCormack & Fortnum, 2013; Abrams & Kihm, 2015). A hearing aid’s frequency-gain response (FGR) is generally fit based on an individual ‘s pure-tone thresholds, and that the fitting process often requires subsequent clinical fine-tuning to address sound quality issues (Anderson et al., 2018). However, clinical fine-tuning has inherent challenges: clinicians cannot directly perceive how each hearing-aid user experiences sound, and hearing-aid users may struggle to articulate their auditory needs in terms of amplification parameters (Caswell-Midwinter & Whitmer, 2021). That is, there are gaps between what a user wants, what information can be conveyed in the clinic, and what a clinician can do with that information.

Allowing users to self-adjust their device’s frequency-gain response (FGR) offers a way to bridge these gaps. Mobile technologies such as smartphones, now a near ubiquitous platform (Ferguson et al., 2026), provide an accessible means for implementing such FGR adjustments. In addition to helping the recall of difficult listening situations (Saunders, 2019), improving patient-provider communication (Convery et al., 2020) and is generally desired by audiologists (Kimball et al., 2018), mobile devices allow wireless-enabled hearing-aid users to make self-adjustments outside the clinic, in the listening situations where adjustments are needed. Self-adjustments can also help overcome challenges such as memory biases, communication barriers, and the issues linked to verifying settings within the controlled acoustics of the clinic. Current hearing-aid self-adjustment apps vary in their interfaces, in the number of controls (e.g., three marked or unmarked sliders), what they control (e.g., filter bandwidths and centre frequencies), the scale of control (e.g., ±6 dB in each filter band), their visual presentation and how immediately accessible they are within the app (e.g., how many button presses it takes for a user to be able to self-adjust their gains). Understanding how users interact with these tools is therefore important, to bolster personalisation in general, to enable environment-specific adjustments that support listening intents, and to serve as input for adaptive technologies designed to deliver automatic adjustments across diverse listening scenarios. Put in a user-experience context (ISO 2019), to promote its use and usefulness in everyday life, an FGR self-adjustment method, like any other user interface, should be easy to understand, effective and efficient.

In terms of ease, previous research in FGR self-adjustment agrees that the number of FGR controls should be restricted to three or less controls to reduce cognitive demand (Dreschler et al., 2008 ; Boothroyd & Mackersie, 2017). In a comparison of different methods with the same underlying filter configuration to change the FGR of speech with different noises, most older participants with mild to moderate hearing loss preferred and were most reliable (test-retest correlation) with the method of adjustment (i.e., two rotary controls for overall gain and spectral tilt) compared to mapping FGR on a two-dimensional surface, pairwise comparisons and categorical loudness scaling (Rennies et al., 2016). In one of the very few studies that have previously compared different self-adjustment control configurations, Dreschler et al. (2008) reported that interfaces that controlled bass and treble, whether dependently or independently, and overall gain were preferred by participants with hearing loss adjusting to audio-visual speech in noise stimuli, and the two-control interface (overall gain and bass/treble tilt) was rated the easiest to use. Compared to hearing-aid personalisation by pairwise comparison (e.g., Zerlin, 1962; Jensen et al., 2019), self-adjustment has potential advantages. Self-adjustment not only allows for multiple comparisons or ‘looks’ between settings, but also, similar to method-of-adjustment matching tasks, permits the use of bracketing: exploring the lower and upper limits of the interval of uncertainty (i.e., the range in which the target and reference are perceived as equal) and then adjusting the target at the centre of the interval, which can increase accuracy (Stevens, 1959) and reduce the bias of the starting point (Wade, 1970).

For self-adjustment to be effective, individuals need to understand how the provided controls affect the sound and how to use them to achieve a desired outcome. Consequently, if these prerequisites are not met, self-adjustments risk being carried out in a trial-and-error manner, potentially increasing the likelihood of failure (Jensen et al., 2019). Evaluating whether individuals self-adjust effectively is challenging, as the chosen settings are guided by inherently subjective and individual criteria. One approach to evaluating the effectiveness of self-adjustment is to assess how consistently individuals can perform self-adjustments using a given method. Several studies have examined the reliability of study-specific self-adjustment procedures, often at a group level, as an indicator of the method’s suitability for any individual using it (e.g., Kuk & Pape, 1992; Elberling & Vejlby Hansen, 1999; Keidser & Alamudi, 2013;Gößwein et al., 2023). Dreschler et al. (2008) found that less reliability (i.e.,greater test-retest standard deviation)for an interface that contained both tilt and mid boost/scoop controls with different step sizes (i.e., multiple controls affect ing the entire spectrum). It is unclear, though, whether the change in reliability was due to overlapping functionality, different step sizes or the particular controls used. Using a two-dimensional surface control (cf. Rennies et al., 2016; Gößwein et al., 2023; Gößwein et al., 2024), Kurşun et al. (2025b) examined how (normalised) FGR adjustments of speech in noise converge over multiple trials, finding that while variance decreased over trials, it was less than ideal for all participants with hearing loss. Nevertheless, it is reasonable to expect that simpler self-adjustment interfaces with fewer, perceptually distinct controls (e.g., a single tilt control; Punch & Robb, 1992)are easier to understand, and alternatively that more complex interfaces with more frequency-band divisions (e.g., the three-band interfaces in many current hearing-aid smartphone applications) allow for more fine-grained and potentially more targeted adjustments. The just-noticeable differences (JND) required to immediately detect a change in FGR increases with narrower frequency bandwidth and increasing centre frequency (Caswell-Midwinter & Whitmer, 2019a/b), and the difference required to elicit a better/worse preference is greater than the JND (Caswell-Midwinter & Whitmer, 2021) and decreases when the duration is increased (Whitmer et al., 2022).

We here conceptualise self-adjustments as aiming – potentially converging – toward an internal reference for sound quality. Hence, we propose that the ease and effectiveness of the method can be psychophysically assessed by replacing this internal reference with an external one, an objective target in a method-of-adjustment task (Fechner, 1860). In these method-of-adjustment tasks, participants typically adjust a single, continuously variable stimulus to match a reference stimulus. In the current context, the measure of interest is the matching error, the signed difference between the adjusted match and the reference, which, when averaged, is used to estimate the point of subjective equality (PSE). The standard deviation of the signed error in method-of-adjustment tasks, a measure of PSE variability, has also been regarded as the JND or difference limen (Gescheider, 1985). JNDs derived in this way have been found to be less, typically by a factor of 2, than those directly measured with other psychophysical methods (e.g. fixed or adaptively tracked same-different pairwise comparisons) in which stimuli are not adjusted by participants (Cardozo, 1965; Wier et al., 1976).By using identical means to have participants match their own self-adjusted FGR as references, we can measure a psychophysical limen for FGR self-adjustment (cf. the JND for the pairwise comparison of FGR; Caswell-Midwinter & Whitmer, 2019a/b) and how it varies with the self - adjustment controls provided to the user.

Different goals in the personalisation of gain, whether inherent to the individual, set by the experimental criteria or suggested by the stimuli used, have led in previous studies to different FGR preferences. Interindividual variability observed in self-adjustment studies may be due to certain character traits that are linked to how they engaged with self-adjustment. Participants ‘readiness’ or ‘anxiousness’ in connection with the self-adjustment method could affect their adjustments (Rennies et al., 2016). While character traits have been previously linked with acceptable noise level (Franklin et al., 2013)and hearing-aid uptake (Cox et al., 2005), neither Perry et al. (2019)nor Walravens et al. (2020) found any relationships between individually preferred FGRs or their consistency, respectively, and various personal characteristics including hearing thresholds and cognitive measures. When participants were instructed to adjust to set criteria, participants on average have preferred more gain for a “speech understanding” compared to a “comfort” criterion (Keidser et al., 2005; Gößwein et al., 2023). When stimuli include speech, there is evidence that speech audibility is an important driver of self-adjustments, even in the absence of instructions to optimise speech intelligibility. Low-level speech (45–55 dB SPL) typically leads to increases in overall gain (Perry & Nelson, 2022) whereas higher level speech (> 65 dB SPL) leads to involve less overall gain than prescribed (Smeds et al., 2006 a/b; Nelson et al., 2018). Individual participants have previously preferred both frequency-gain increases and decreases to supra-threshold stimuli (Nelson et al., 2018; Gößwein et al., 2023; Kliesch et al., 2023), consistent with each individual weighing multiple criteria and seeking a best-possible compromise between them (e.g., speech audibility and loudness comfort). Self-adjustments to speech-in-noise stimuli in which noises dominated one part of the spectrum have resulted in reduced gain in that part of the spectrum, essentially improving SNR (Keidser et al., 200).5 In contrast, Gößwein et al. (2023) did not observe an effect of stimulus on the average preferred frequency-gain slope, irrespective of adjustment criterion, possibly because all the noises were relatively broadband. Speech in noise, especially at less-adverse SNRs, has produced more reliable self-adjustments compared to speech in quiet (Keidser et al., 2005; Nelson et al., 2018). Only D’Onofrio et al. (2019) included music stimuli, but they did not observe any differences due to stimuli. Given that participants may naturally change how they weigh and apply their own adjustment criteria with the content of stimuli, it would be informative to assess the effect of interface without setting a criterion and the effect of stimulus content (speech in quiet, in noise or music) while providing spectrally similar stimuli. To be efficient, the method should also be quick while still effective, allowing the user to adjust the FGR to the scenario while the scenario– and the current goal in self-adjustment – is still relevant. Early studies of self-adjustment described multiple spectraltilt procedures taking 10 minutes or more (Punch & Robb, 1992), whereas more recent studies have described adjustments taking approx. 1 minute (Rennies et al., 2016; Kliesch et al., 2023) . An optimal solution for self - adjustment is then an interface that balances ease and efficiency – simplicity and speed – with effectiveness– perceptual relevance and repeatability.

The current study examines how the individual, the interface and the stimulus influence the ease, effectiveness and efficiency of hearing-aid gain self-adjustment. Specifically, we compare three self-adjustment interfaces that differ in the number and underlying filter configuration of their control(s), thereby varying the flexibility in FGR adjustment they afford: (1) a single control for adjusting spectral tilt changing bass and treble jointly in opposite directions (e.g., Punch & Robb, 1992), (2) two controls for adjusting bass and treble independently (e.g., Boothroyd & Mackersie, 2017), and (3) three controls for adjusting bass, mid and treble (e.g., Dreschler et al., 2008). Across three stimulus types, we examine the reliability of self-adjusted gain settings (cf. Kurşun et al., 2025b), interaction behaviours (cf. Gößwein et al., 2024), and whether there are links between interaction behaviours and self-adjusted gain . To better understand the trade-offs between ease and effectiveness, we also examine how participants rated each adjusted sound(cf. Sabin et al., 2020) and which interface they preferred(cf. Dreschler et al., 2008 ; Rennies et al,. 2016). The study also explores whether demographic, hearing-related, and personal characteristics help explain individual differences in preferences (cf. Perry et al., 2019; Walravens et al., 2020) and whether there are any patterns in those preferences across individuals (cf. Balling et al., 2021). Additionally, within the same participants, the current study examines whether a similar sound-matching task can serve as an objective measure to evaluate effectiveness, an individual’s ability to interact with self-adjustment interfaces in a meaningful way, and to what extent interaction behaviours are shared across the self-adjustment and sound-matching tasks. By examining these different elements, we aim to answer how the method, the signal and the individual matter in the self-adjustment of hearing-aid gain.

## METHODS

### Participants

Forty-one bilateral hearing-aid users (16 female) participated in the self-adjustment experiment. The median age was 73 years (range 61–82). All participants were initially identified from patients and referrals to local hearing clinics (NHS Greater Glasgow & Clyde). Recruitment and all procedures were approved by the West of Scotland Research Ethics Committee (18/WS/0007) and NHS R&D (GN18EN094). All participants’ ear canals and eardrums were verified to be healthy and free of blockage by otoscopic inspection prior to air - and bone-conduction pure-tone audiometry conducted on the same day as the self-adjustment task. All participants had sensorineural hearing loss with air-bone gaps < 15 dB (based on averages between 0.5–4 kHz). The left panel of Figure 1 show individual pure-tone thresholds. The median better-ear four-frequency (0.5, 1, 2, and 4 kHz) average (BE4FA) was 38.75 dB HL(range 18.75–56.25dB HL). Left-right asymmetry in four-frequency averages ranged from 0-11.25dB HL (median 2.5 dB HL). BE4FA was not correlated with age (r = 0.02; *p* ≫ 0.05). Each participant’s individual better-ear NAL-R gain prescription (Byrne & Dillon, 1986) served as a starting point from which adjustments were made in the self-adjustment task(middle panel of Figure 1). NAL-R was used because the formula would potentially represent an individual starting point that was not necessarily ideal compared to current prescriptive formulae or similar to participants’ current settings.

**Figure 1.**
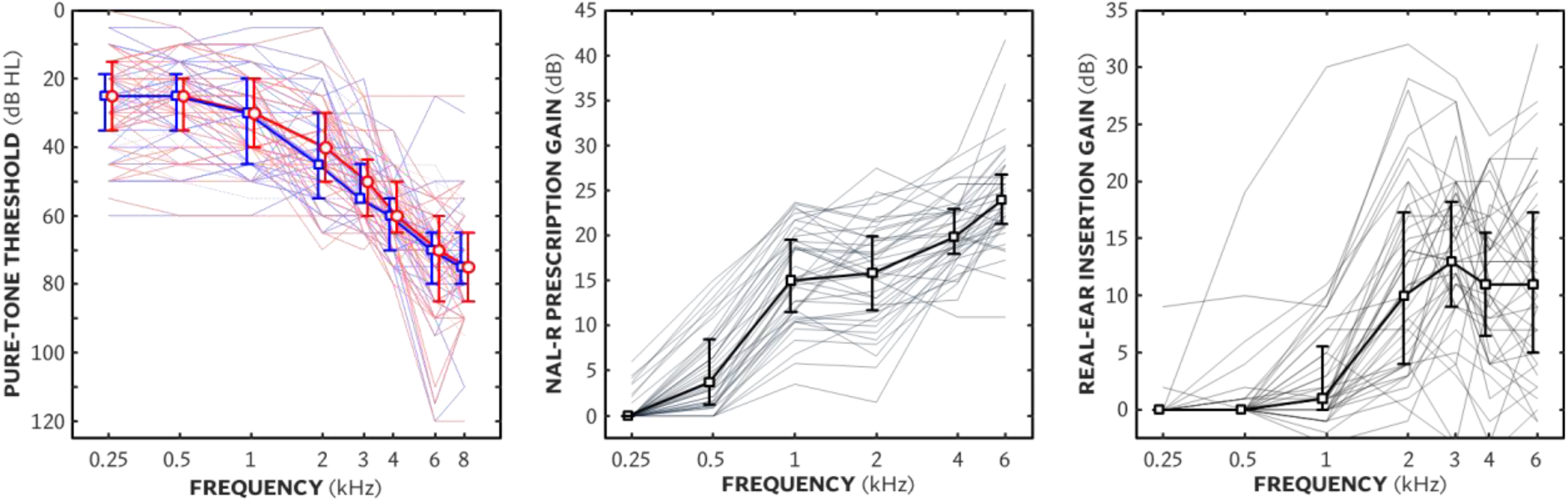
Left panel shows individual (thin lines) and median (bold lines) air-conduction pure-tone thresholds as a function of frequency for left (blue, median squares) and right (red, median circles) ears. Error bars show interquartile range. Middle panel shows individual and median NAL-R prescription gain as a function of frequency for each each participant’s better ear. Right panel shows individual and median rea-lear insertion gain as a function of frequency.

In a second session that occurred more than four weeks (29–153 days)after the first session, 36 of the 41 participants returned to complete several additional tasks including a sound-matching task. During the sound-matching task, two participants wore their hearing aids during testing, and two other participants did not comply with instructions during testing . Of note, all four were compliant during the self-adjustment task in their previous session. These participants’ data was excluded from the sound-matching task, resulting in data from 32 participants (12 female). The median and range of ages and BE4FAs did not change from the first session (reported above).

Median hearing-aid use was 7 years (range 1-28 years). Most participants (35) currently wore Resound/Danalogic behind-the-ear (BTE) devices, four with standard tubing and earmoulds, the remainder with thin tubes and domes, that were fit by NHS Greater Glasgow and Clyde audiology clinics. Of the remaining six participants, two wore completely in the canal devices, and four wore receiver-in-canal BTE devices. Real-ear insertion gain (REIG) for each device was measured for 65 dB using the ISTS (International Speech Test Signal; Holube et al., 2010). All probe-tube measurements of REIG were conducted using Affinity 2.0 (Interacoustics) and with the volume controls and programmes set as participants would most regularly set them if at all. The REIGs of each participant’s better-ear are shown in the right panel of Figure 1.

### Apparatus

Participants sat in a sound-dampened, double-walled audiometric booth (IAC Acoustics)with a 23.6-inch touchscreen (iiyama ProLite T2435 MSC) in front of them. Participants listened through circumaural open-back headphones (AKG K702). Headphones were equalised to within ±4 dB from 0.08-8 kHz in 1/3-octave bands. A Babyface Pro (RME) provided digital-to-analogue conversion for the stimuli manipulated in MATLAB, and a headphone amplifier (Samson S-Phone) provided additional headroom to minimise clipping with maximum gain adjustment in any band.

### Stimuli

Three stimuli were used: (1) speech in quiet, (2) speech in noise and (3) music. *Speech in quiet* stimuli consisted of segments a male talker with central Scottish accent reading *The Musgrave Rital* by Sir Arthur Conan Doyle (Macpherson & Akeroyd, 2013). *Speech in noise* stimuli consisted of different segments from the same speech recording in cafeteria babble noise (Bjerg & Larsen, 2006) at +5 dB SNR RMS. *Music* stimuli were segments from Barry White’s “I Can’t Get Enough Of Your Love Babe” (White, 1974). Original stimuli were recorded (for the music, remastered) at 16-bit and 44.1 kHz sample rate. Fourteen unique segments for each stimulus type were used: two for practice trials, and 12 for experiment trials. Stimulus durations ranged from 14–21 s (mean 17 s)to maintain the varying ends of speech and musical phrases as stimuli playback was continuously looped until participants paused or proceeded to the next trial. Within each stimulus type, the power spectrum of the 12 test segments varied (σ) by 1.1-1.2 dB averaged across third-octave bands. All stimuli were gated with 25-ms raised-cosine onset and offset ramps. NAL-R gain was applied using an 140-tap finite impulse response filter to 24-kHz resampled stimuli (Kates & Arehart, 2010). Filtered stimuli were then resampled to 44.1 kHz and 32-bit resolution to allow for a sufficiently large dynamic range during stimulus playback. Headphone levels were calibrated to give a long-term A-weighted average of 65 dB SPL (with a NAL-R response of 0 dB gain and without adjustable gain applied), which was verified using an artificial ear with a level meter (Brüel & Kjær Artificial Ear 4152 & Sound Level Meter 2260). The same equipment was used to equalise the headphones and verify the function of each filter in each interface (responses using white noise as input are shown in Figure 2).

**Figure 2.**
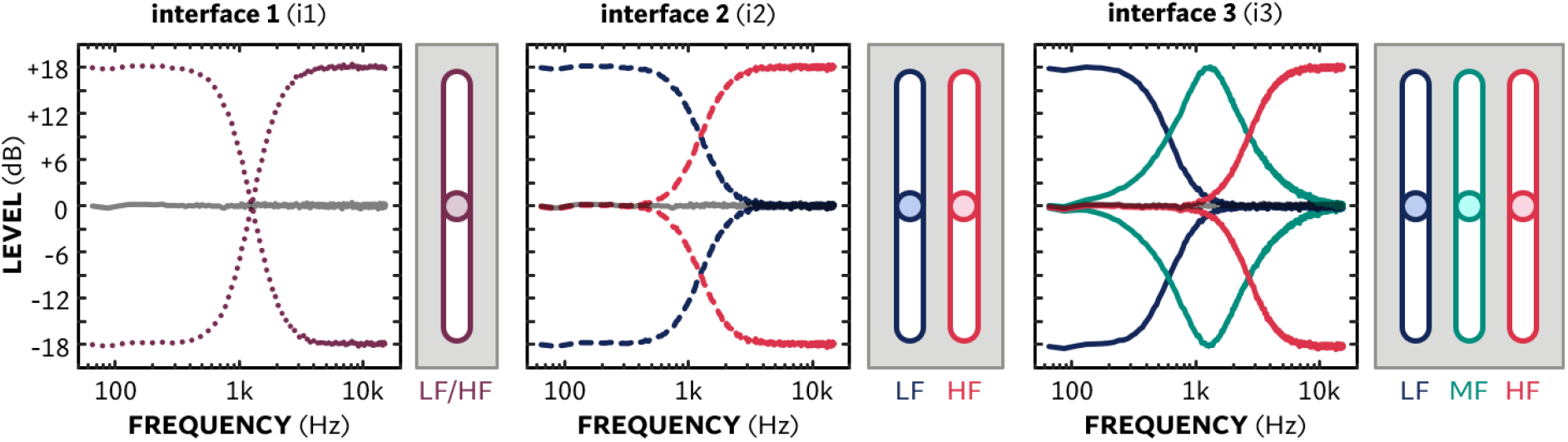
Schematics including measured filter responses es of the three interfaces illustrating the±18-dB adjustable range as a function of frequency for each interface. Left panels shows interface 1(i1), a single slider adjusting LF and HF gain in opposite directions. Middle panels shows interface 2(i2), two independent sliders for LF and HF Right panels shows interface 3(i3), three independent sliders for LF, MF and HF.

### Interfaces & adjustable gains

There were three different control configurations with one, two and three unlabelled vertical sliders (see Figure 2). The slider order in interfaces 2 and 3 was fixed and all sliders provided gain of ±18 dB from the central neutral position (i.e., a total range of 36 dB). Interface 1(i1) consisted of a single vertical slider jointly controlling the gains of low and, inversely, high-shelf filters, each with a cut-off frequency (filter response-3 dB re peak) of 1250 Hz. Interface 2 consisted of two vertical sliders independently controlling the gains of low-shelf (left slider) and high-shelf (right slider) filters with cut-off frequencies of 1250 Hz.Interface 3 consisted of three vertical sliders, corresponding to a low-shelf filter (left slider) with cut-off frequency of 600 Hz, a peaking filter (middle slider) with centre frequency of 1250 Hz and high-shelf filter (right slider) with cut-off frequency of 2600 Hz (i.e., a bass-mid-treble configuration). In addition to the sliders, each interface included a *Play*/*Pause* toggle and *Submit* buttons.

The filters controlled via the interfaces were implemented using the *multibandParametricEQ* function (MATLAB Audio Toolbox), which uses 2nd order IIR filters. The toolbox’s shelving filters were limited to a maximum gain of ±12 dB. To allow the gain range of ±18 dB per slider, two filters (with controllable maxima of 12 and 6 dB) with the same specifications were put in series. Coefficients for the slopes of the low and high-self filters were both set to 1 and peaking filter’s Q was set to 0.6 to obtain a flat spectrum (±1 dB in 1/3 octave bands from 0.1 –8 kHz) when the gain in all filters was increased or decreased by the same amount. Slider resolution (i.e., the smallest change in gain possible) was 0.036 dB.

### Questionnaires

Participants completed four questionnaires in the same session as the self-adjustment task: the International Outcome Inventory for Hearing Aids (IOI-HA) and Technology Readiness Index (TRI) prior to self-adjustment, the Big Five Personality Index (BFI) between repeats 2 and 3 (i.e., in the middle of the self-adjustment task), and the Locus of Control Scale (LOC) following self-adjustment. The IOI-HA (Cox & Alexander, 2002) assesses hearing-aid benefit using seven items, each representing a different domainwith a discrete 5-point scale; the outcome is reported as the sum score of all questions (possible range 7–35). The TRI (Parasuraman et al., 2015) contains ten items with 5-point Likert-type responses split into three subscales assessing attitudes, values and impact of technology. The version of the BFI used here (John et al., 1991; Johnet al., 2008) contains 44 items with a 5-point Likert scale. divided into five subscales associated with openness, conscientiousness, extroversion, agreeableness and neuroticism. Answers are given on a 5-point Likert scale. The LOC questionnaire (Levenson, 1974) contains 24 items with a discrete 6-point scale of agreement without a neutral (0) midpoint divided into three subscales assessing how much one’s circumstances are controlled by chance, external or internal forces. Participants were also asked if they had experience adjusting audio (e.g., the bass and treble controls on a car radio, home stereo or TV) or video (e.g., changing the image brightness, colour balance, or sharpness on a TV). After completing the self - adjustment task, before debriefing, participants were asked to choose which interface they preferred and to briefly describe what influenced their choice.

### Self-adjustment procedure

Seated in the audiometric booth, the self-adjustment to preference task was explained to the participants verbally and also provided during the task in written form. On each trial, they were instructed to adjust the sound “to your preference” with one of the three interfaces and stimuli described above. Each trial started with all sliders set to the centre of the adjustable range. Participants could then adjust the sliders without time limitations while listening to a continuously looped stimulus. After completing each self-adjustment by tapping the *Submit* button, the final adjusted sound kept playing while a window appeared in which participants were asked to “please rate the adjusted sound” by selecting 1–5 stars. After a pause of 200 ms, the next trial started.

The self-adjustment task started with six practice trials that were chosen from the combinations of interfaces and stimulus types to include at least two instances of each interface and each stimulus type. Participants could ask questions or ask for help or clarification during and after the practice trials. Per stimulus type, two unique segments were used for the practice trials, and the remaining 12 unique segments of each of the three stimulus types were presented once during the experimental trials. The 36 experiment trials were organised into four blocks, each block comprised of the nine unique combinations of stimulus type and interface type, which were presented in a pseudo-random order using incomplete Latin squares. There was a break after the first two blocks (18 trials) in which participants completed the BFI questionnaire.

### Sound-matching procedure

In each sound-matching trial, participants listened to two versions of the same sound sample that differed only in FGR. They were instructed to adjust one of the sounds – hereafter referred to as the *match sound*– to match the other – hereafter referred to as the *reference sound*– using the same interfaces they used during the self-adjustment to preference task. Participants could freely switch between the match and reference sounds, which were continuously looped, to compare them as often as desired to find a match. The 36 sound-matching trials corresponded one-to-one to the self-adjustment trials, reusing the endpoints – the preferred gains – for each stimulus segment and interface as the reference FGRs for sound-matching, with the trial order randomised . The sound - matching task included 6 practice trials, followed by the 36 data-collection trials, with a break after the first 18.The initial FGR of the match sound was obtained by applying a randomly selected positive (+) or negative (-) gain adjustment in the continuous range of 5 –12 dB per slider. For the four repetitions in each stimulus category, the sign of initial offset for sliders in each interface was balanced for i1 and i2 and always included an opposite offset for i3.

### Data analysis

Analyses were conducted in MATLAB (version 2021b) and IBM SPSS Statistics (version 29.0.1.0). To determine the appropriate analyses, the normality of the data distributions was evaluated using Shapiro-Wilk tests (Shapiro & Wilk, 1965), visual inspection of Q-Q plots, and consideration of outliers defined as values exceeding 1.5 times the interquartile range (IQR). Parametric tests were used when the data were found to be approximately normally distributed; otherwise, the corresponding non-parametric tests were applied. For parametric data that lacked sphericity, the degrees of freedom were adjusted using the Greenhouse-Geiser correction. Based on these criteria, only the self-adjusted endpoints were considered normally distributed. The appropriate tests for each dataset are indicated at the outset of each analysis within the Results. Unless otherwise indicated, all correlational analyses were rank correlations reporting Spearman’s rho (ρ). For multiple comparisons involving the same data, *p* values were adjusted using the Bonferroni correction(Dunn, 1961). Given how bandwidth and centre frequency affect gain discrimination (Caswell-Midwinter & Whitmer 2019a), we initially treated the sliders in interfaces with multiple sliders separately for all measures (i.e., comparing the six sliders, not the three interfaces) except for trial durations, sound ratings and interface preferences.

For pattern analysis in the self-adjustment data, k-means cluster analysis was used (Steinhaus, 1956). This method was chosen for the availability of established approaches to determine the optimal number of clusters: (a) the gap statistic (Tibshirani et a l., 2001), (b) the silhouette test (Rousseeuw,1987), which identifies the number of clusters with the highest silhouette coefficient, and (c) the elbow method (Thorndike, 1953), which compares the squared distance of each data point to the cluster centre. As the self-adjustment data were expressed on the same scale as deviations from an initial starting point, no normalisation or scaling was applied in the cluster analysis. Cluster stability was assessed by calculating the agreement between (a) clusters produced by k-means and partitioning-around-medioids methods (Kaufman & Rousseeuw, 1990) and (b) the random removal of participants from the dataset (Hubert & Arabie, 1985; Hennig, 2007).

## RESULTS

### Self-adjustment

#### Self-adjustment endpoints

For each slider in each interface with each stimulus, participants’ mean self-adjustment endpoints from the four repeated adjustments were calculated as the gain (in dB) relative to the initial NAL-R gain; results are shown in the left panel of Figure 3. The effect of stimulus and slider on self-adjustment endpoints was assessed with a two-way 3 × 6 repeated-measures ANOVA. There was a significant main effect of stimulus type [*F*_(2, 80)_ = 10.51; *p* < 0.001; η^2^ = 0.21]; the average gain for speech in noise was 1.2 and 0.7 dB less than music and speech in quiet, respectively (paired Student’s *t*_(40)_ = - 4.41 & -2.98; *p* < 0.001 & *p =* 0.0048, respectively). There was also a significant main effect of slider [*F*_(1.77, 70.74)_ = 40.26; *p* < 0.001; η^2^ = 0.50]. Post-hoc paired Student’s *t*-tests showed that on average i1LF gain was greater than any other slider (Δ = 4.7-10.9 dB; *t*_(40)_ = 5.34-8.27; all *p* < 0.001), average i2LF gain was less than i3LF (Δ = -1.2dB; *t*_(40)_ = -4.44; *p* = 0.001), and i2HF gain was less than i3HF gain (Δ = -1.2 dB;*t*_(40)_ = -3.45; *p* = 0.020). Within interfaces, the low-frequency gains (i2LF and i3LF) were significantly greater than the higher frequency gains (i2HF, i3MF and i3HFΔ; = 3.8-5.5 dB; *t*_(40)_ = 4.02-5.17; all *p* < 0.05). The interaction of slider and stimulus was also significant [*F*_(7.05, 281.86)_ = 2.23; *p* = 0.032, η^2^ = 0.053]; gains involving low and mid-frequencies were greater for music than speech-in-noise (Δ = 1.2-2.1dB; *t*_(40)_ = 2.53-4.19; all *p* < 0.05).

**Figure 3.**
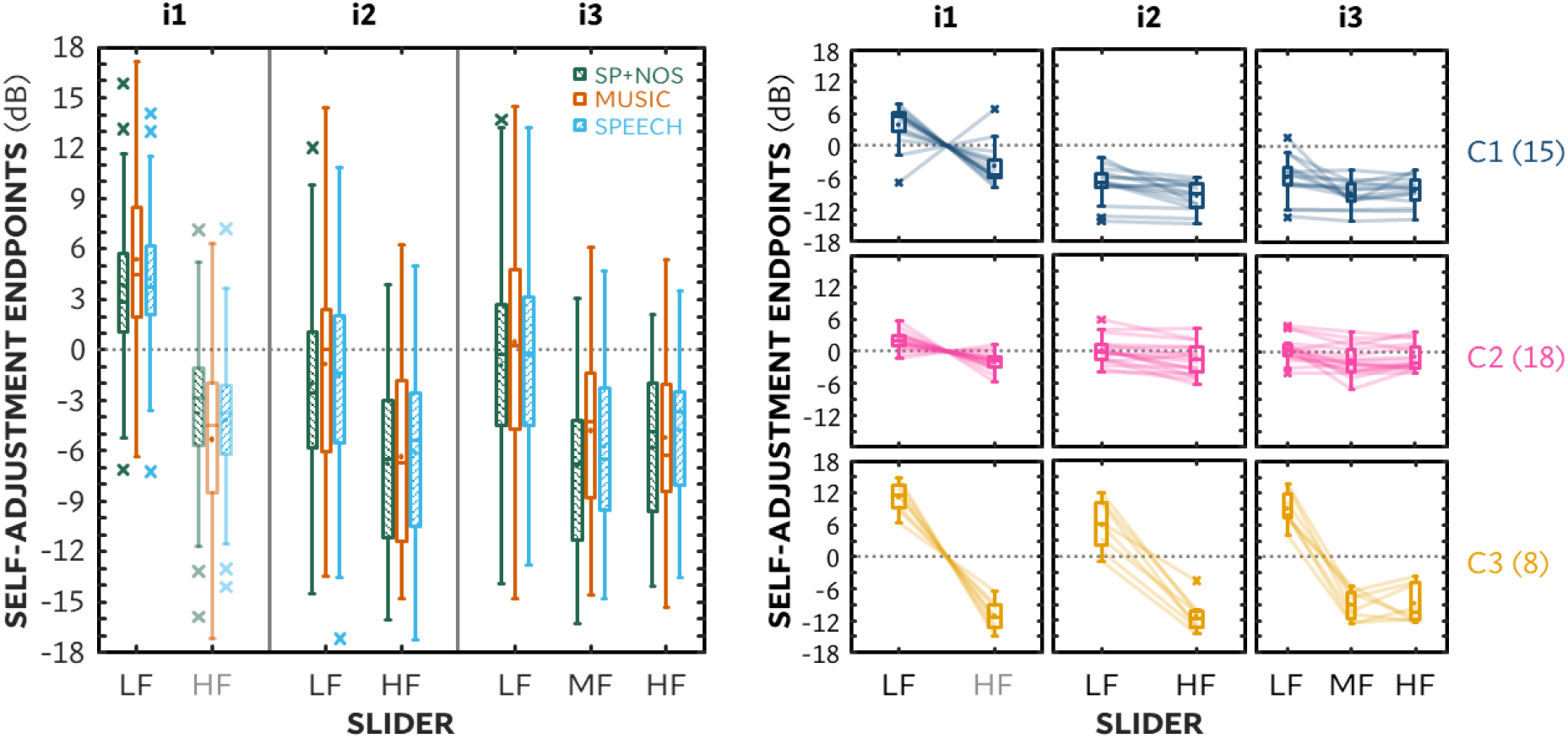
Mean self-adjustment endpoints (i.e., preferred gain setting away from NAL-R prescription)as a function of interface and slider. Left panel shows endpoints for the three stimulus s types averaged across four triasl:speech in noise (SP+NOS), music and speech (in quiet). Right panel shows endpoints for the three three clusters averaged across 12 trials 4(repeats × 3stimulus types) both as individual lines and boxplots. Note that for i1 in both panel, sHF end points are the inverse of the corresponding LF endpoints. Horizontal line within boxes = medians, small circles = means;boxes= interquartile range(IQR); whiskers= data within 1.5 × IQR; outliers< 1.5 × IQR = crosses.

Outwith the large group mean differences in gain set with i1 compared to the gains set with i2 and i3, these group mean gain differences were generally small, in the order of1–2 dB. Counter to these small differences in endpoint means, however, was the large interindividual variation, with the range of endpoints spanning 16.2-29.3 dB. To see if there was any relationship between the variance in these self-adjusted gains and individual current hearing-aid gain which also varied substantially above 250 Hz (right panel of Figure 1), we compared these endpoints, representing differences between preferred gains and NAL-R prescriptions, to differences between current hearing-aid REIGs and NAL-R prescriptions. Individual self-adjusted gains with any slider in the current study were not significantly correlated with the difference between REIG and NAL-R gains at any corresponding octave-band centre frequency (Pearson product-moment correlation coefficient *r*^2^ = 0.0019-0.030; all *p* >> 0.05) . To probe if experience with hearing aids might bias self-adjusted gains towards current hearing-aid gains, we split the participants into those with less and more than 5 years of hearing-aid use (*n* = 17 & 24, respectively). There were no significant differences in post-hoc comparisons of less and more experienced users at any centre frequency for either self-adjusted endpoints (Student’s *t*_(39)_ = 0.0037-0.86; all *p* > 0.05) or the difference between those endpoints and current hearing-aid gain (*t*_(39)_ = 0.089-0.56; all *p* > 0.05).

#### Clustering by self-adjustment endpoints

To explore patterns in self-adjustment endpoints and to evaluate their relationship to participant characteristics, self-adjustment endpoints were grouped employing k-mean cluster analysis. As most stimulus effects were in the range of-21 dB,cluster analysis used the self-adjustment endpoints per slider (i1LF, i2LF, i2HF, i3LF, i3MF and i3HF) averaged across stimuli. The optimal number of clusters was determined using the Silhouette plot, the elbow method and the gap statistic (see Data Analysis section above) . The first two methods agreed on three clusters ; the third (gap statistic) returned one cluster as optimal followed by three as second best. Based on these results, three clusters were chosen Figure 3 (right panel) shows the endpoints for each cluster and Table 1 summarises the endpoint cluster centres . The most pronounced differences between clusters were for independent low-frequency adjustments; the three clusters differed by means of 6 dB or more for both i2LF and i3LF.

**Table 1.**
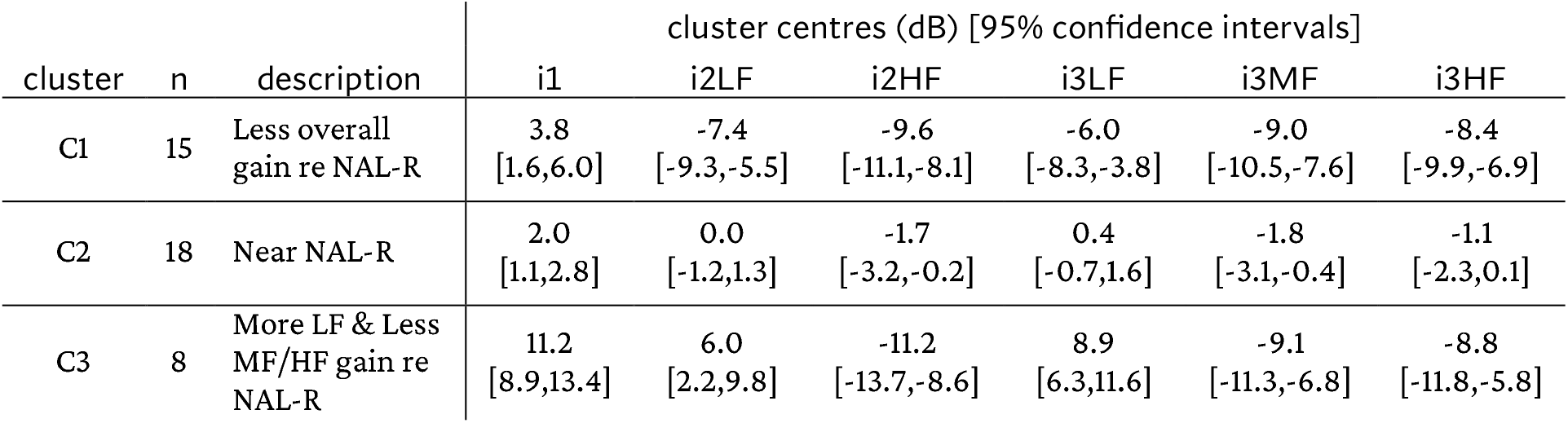
Cluster size, description of its characteristics, cluster centres (means) per slider with 95% confidence intervals in brackets based on self-adjustment endpoints averaged across repetitions and stimuli.

**Table 2.**
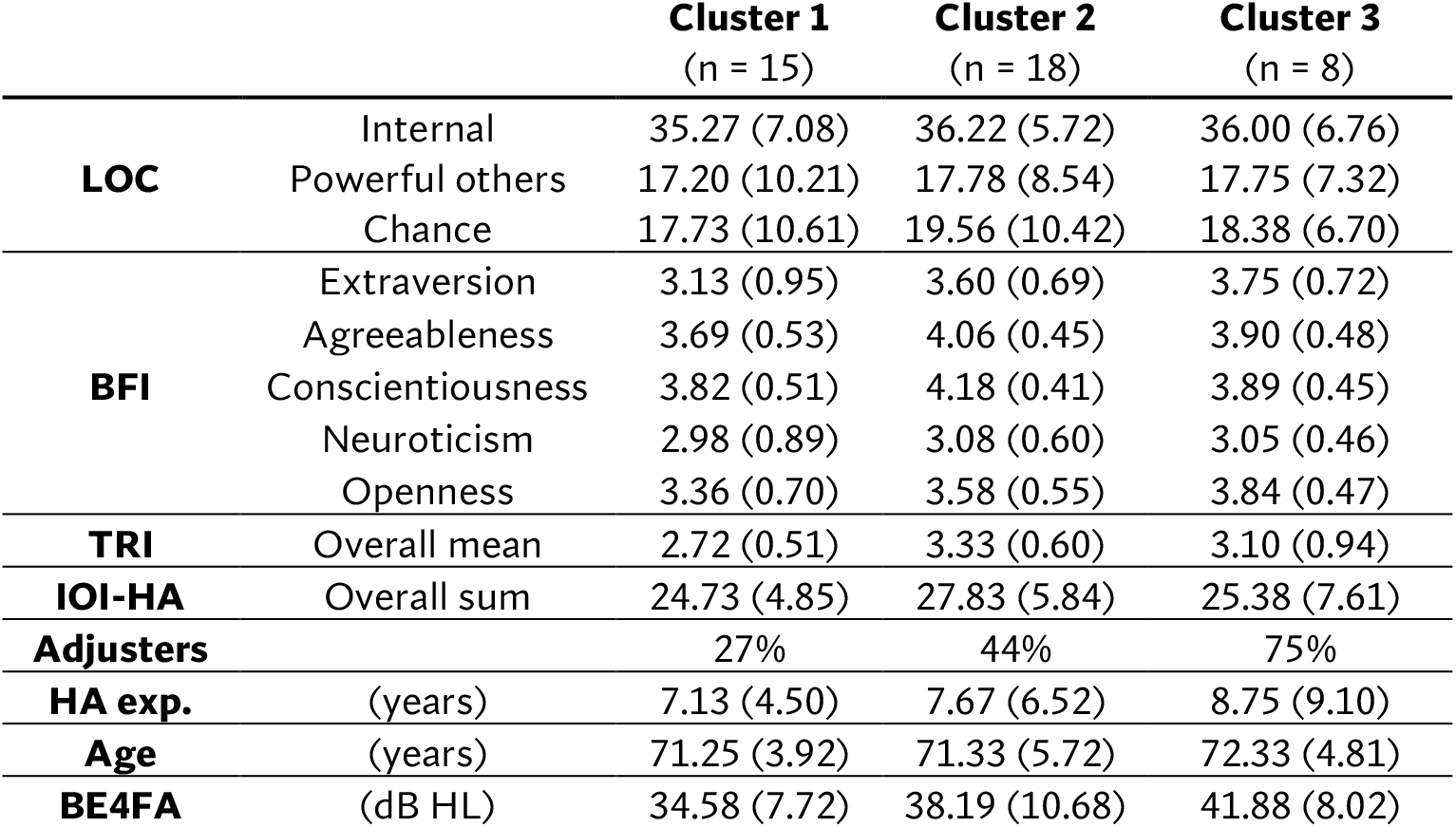
Means and standard deviations (in parentheses) of questionnaire scores, hearing-aid (HA) experience, age and better-ear four-frequency pure-tone average thresholds (BE4FA) for each cluster. See Methods for further information on the questionnaires.

To test the robustness of the clusters, the results of two clustering algorithms were compared: k-means and partitioning-around-medioids. The adjusted Rand index, a coefficient of agreement (0-1 range)comparing cluster affiliation with chance-agreement adjustment (Hubert & Arabie, 1985)was 0.93, indicating that the cluster solution was robust to the choice of clustering algorithm. To assess the stability of the clustering, the k-means analysis was repeated 1000 times with participants randomly removed and agreement between each new clustering and the original solution quantified using the mean adjusted Rand index. Removing eight participants (19%; n.b., the number of participants in C3) produced a mean adjusted Rand index of 0.99, and 89% of the original affiliations were reproduced, confirming that the clusters were generally stable given the sample size (cf. Hennig, 2007).

Table summarises participant characteristics, including age, BE4FA and questionnaire scores, by cluster. We tested for differences in these participant characteristics between clusters using Kruskal-Wallis ANOVA for all continuous variables . There were no significant differences between clusters for any participant characteristics [χ^2^(2,38) = 0.023-3.98; all *p* > 0.05] . The majority of C3 participants were “adjusters”, defined as those that reported previous experience adjusting either sound or video equipment. However, there were still more”adjusters” in the larger cluster C2 than C3 (8 vs. 6, respectively). The association between being an adjuster and cluster was statistically assessed using Pearson’s χ^2^ test of the adjuster × cluster contingency table ; there was no t a significant association [χ^2^_(2,38)_= 4.95; *p* > 0.05]. We also compared clusters using separate Kruskal-Wallis tests for REIGs at any corresponding octave-band centre frequency from 250-4000 Hz . There were no significant REIG differences between the three clusters [χ^2^_(2, 38)_ = 0.50-4.59; all *p* > 0.05] nor for the differences between REIG and NAL-R gains [χ^2^_(2,38)_ = 0.33-1.19; all *p* > 0.05].

#### Reliability of self-adjustments

Participants adjusted gains four times for each interface and stimulus type. Self-adjustment reliability was calculated as the standard deviation of the four endpoints of each slider and stimulus type. Note that for each repetition of a stimulus, the participant heard a different segment from the same stimulus type that was spectrotemporally similar to the other segments. Given the skew in the data, the effect of stimulus and slider on reliability were tested with Friedman two-way ANOVA by ranks tests. There was no significant main effect of stimulus type on reliability [χ^2^_(2, 80)_ = 1.02;*p* = 0.60], hence reliability was pooled across stimulus types as well as repetitions for each slider(i.e., individual slider reliabilities were the standard deviation of the 12 endpoints, four repeats of the three stimulus types). Figure 4 shows the reliability results across all participants, independent of cluster. The red dotted line indicates the mean standard deviation for random adjustments (9.8 dB). The overall median reliability was 2.8 dB, although the range varied across individuals from1.0-5.5 dB. There was a significant main effect of slider on reliability [χ^2^_(5, 200)_ = 14.17; *p* = 0.015; Kendall’s *W* = 0.01], but the only significant difference revealed in Wilcoxon rank-sum post-hoc comparisons was very small: i2HF was 0.4 dB more reliable than i3HF (*z* = -3.05; *p* = 0.034). There were no significant differences between clusters in reliability, assessed as the median standard deviation across sliders using the Kruskal-Wallis ANOVA [χ^2^_(2, 38)_ = 4.18; *p* = 0.12]. Additionally, we probed procedural effects using Friedman’s test. Reliability averaged across sliders and stimuli did not significantly vary with repeated blocks [χ^2^_(3, 120)_ = 7.10; *p* = 0.069].

**Figure 4.**
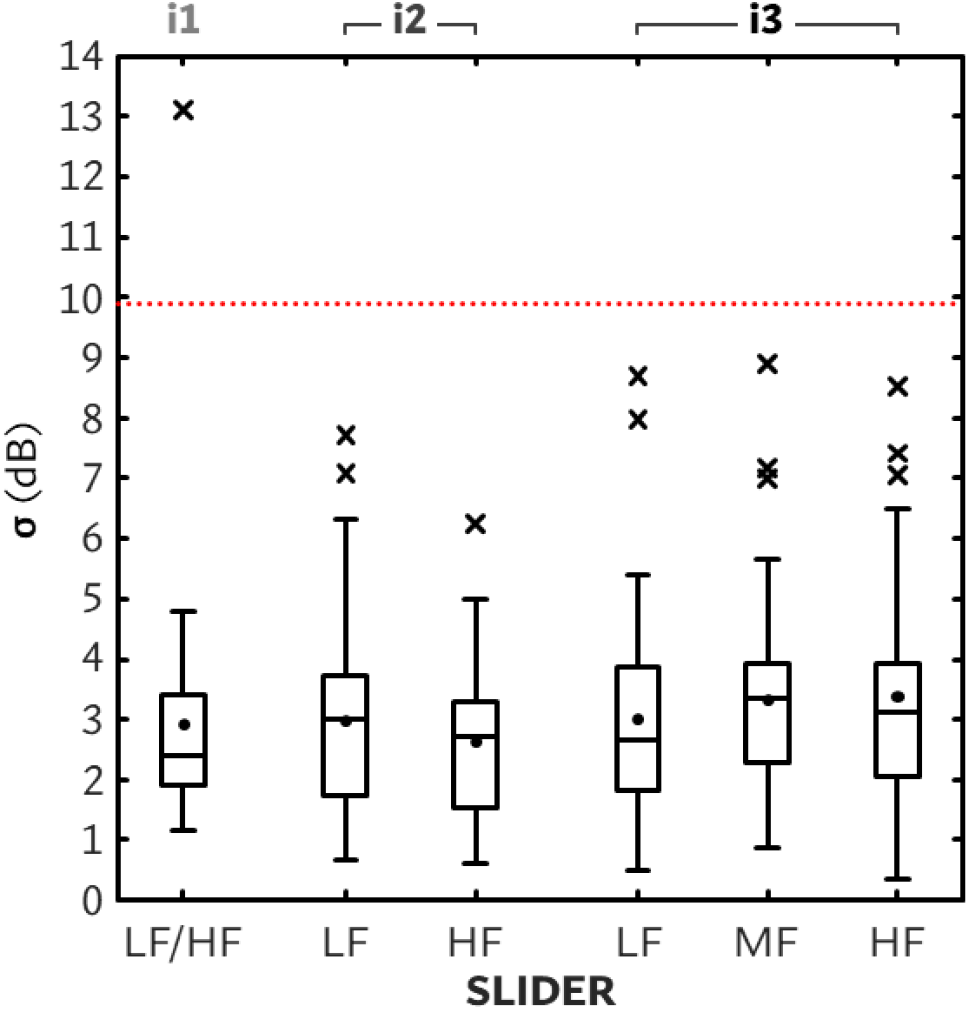
Standard deviations of self-adjusted gain endpoints as a function of interface (indicated at top of panel) panel) and slider. The red dotted line indicates the mean standard deviation for random adjustments. For description of boxplot symbols, please refer to the caption of Figure 3.

#### Self-adjustment behaviours

Three basic behavioural metrics were analysed from participants’ adjustments: trial duration, ranges explored and reversal rate . Trial duration, the most direct measure of efficiency, was calculated as the time elapsed from the start of a trial until participants submitted their self-adjustment endpoints (i.e., the time spent both adjusting and appraising). The range of gains explored per slider has the potential to reveal differences in interaction behaviour, such as bracketing, as well as perception, such as detecting a change. The range was calculated as the difference between maximum and minimum slider positions for each trial. Reversal rate, another potential indicator of uncertainty in adjustment (cf. head rotation reversals in orientation tasks; e.g., g., Whitmer, McSheffery et al., 2022), was calculated from the sum number of slider direction changes, independent of other actions (e.g., if a participant adjusted a slider up, then adjusted another slider, then returned to the same slider and adjusted down, that was counted as a reversal), made within each trial divided by the trial duration. Examples of slider adjustments with each interface (columns) for a participant in each of the three clusters (rows) are shown in Figure 5A. Examples were chosen as archetypes of different behaviour patterns as well as being representative of relative endpoints for each cluster . The adjustment duration can be seen to increase with interface for the C2 participant (middle row) whereas it appears to decrease for the C1 participant with i3. The range explored by the C3 participant in Figure 5A covers the extremes, whereas the range of the C2 participant’s adjustments was relatively small. Reversal rates are shown to encompass both bracketing-like behaviour and smaller adjustments, such as the C3 participant with i1 converging towards an endpoint (cf. Kurşun et al., 2025b) with near maximum LF gain (bottom right panel) . Figure 5B compares these time-based interaction behaviours within participants: trial durations between interfaces (top row), ranges between sliders controlling similar frequency regions (middle row) and reversal rates between interfaces (bottom row). Given the visible positive skew in these behavioural measures, the effects of slider and stimulus on these measures were assessed with Friedman two-way ANOVA by ranks tests followed by Wilcoxon signed-rank post-hoc comparisons; differences in clusters were assessed with Kruskal-Wallis one-way ANOVA tests followed by Wilcoxon rank-sum tests.

**Figure 5A.**
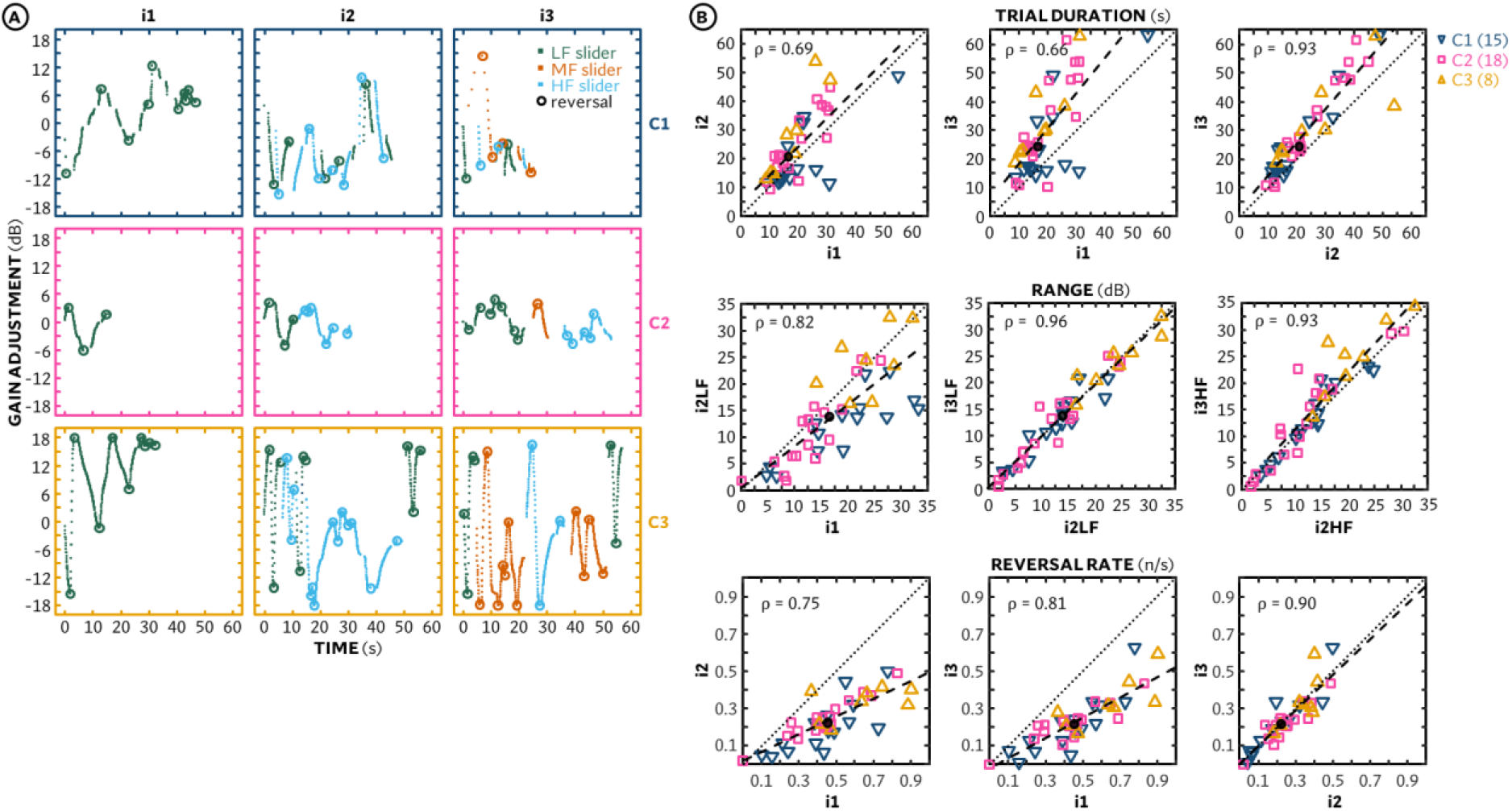
Panels show examples of adjustment behaviour for each interface (columns) from a participant from each cluster (rows); different slider activity is indicated by colour [dark green = low-frequency (LF); orange = mid-frequency (MF); blue=high-frequency (HF) / and reversals by open circles. Figure SB. Top right panels show individual trial durations in seconds, averaged across stimulus types, comparing each interface with each other. Middle panels show range of adjustments in dB, averaged across stimulus types, comparing frequency-band relevant sliders across interfaces. Bottom panels show reversal rates, averaged across stimulus types, comparing each interface with each other. Symbols indicate cluster, determined by average endpoint [dark blue downward triangles = cluster 1 (Cl); pink squares = cluster 2 (C2);gold upward triangles = cluster 3 (C3)]. Dashed lines show linear regressions of the data; black filled circles show medics.s

##### Trial duration

There was a main effect of interface on trial duration [χ^2^_(2, 80)_ = 37.90; *p* < 0.001; Kendall’s *W* = 0.46]; durations increased linearly by approx. 4 s with the number of sliders per interface, from a median of 16.5 s for i1 to 24.3 s for i3 [*z* = 3.22-4.85; all *p* < 0.01]. There was an effect of stimulus type on trial duration [χ^2^_(2, 80)_ = 17.90;*p* < 0.001; Kendall’s *W* = 0.22]; speech in noise trials took approx. 2 and 3 s longer to complete than music and speech trials, respectively (*z* = 3.56 & 4.04; both *p* < 0.01). However, the variation amongst individuals, as shown in the top panels of Figure 5B, was much larger than interface or stimulus effects, ranging from 10-55 s and was significantly correlated across interfaces (Spearman’s rank-correlation coefficient *ρ* = 0. 66–0.93; all *p* ≪ 0.001). There was also a significant effect of repeat (i.e., trial-block number) on median trial duration [χ^2^_(3, 120)_ = 11.84;*p* = 0.0080] ; trials in the first two blocks took 1.9s longer than the last two blocks to complete (*z* = 2.83; *p* = 0.0046) .

##### Range explored

There was a significant main effect of slider on range explored [χ^2^_(5,200)_ = 26.64; *p* ≪ 0.001; Kendall’s *W* = 0.13]. The range explored with the i1 slider was significantly greater than all other sliders save for i3HF (median Δ = 2.6-4.1dB; *z* = 3.55-4.26; all *p* < 0.01), and the range with i3HF was greater than with i3MF (median Δ = 3.2 dB; *z* = 2.85; *p* = 0.026). Ranges also significantly differed as a function of stimulus type [χ^2^_(2,80)_ = 18.20; *p* < 0.001; Kendall’s *W* = 0.11], with a smaller range explored for speech than for speech in noise and music (median Δ = -3.7 & -0.8 dB, respectively; *z* = - 4.84 & -2.79; both *p* < 0.05). These differences, however, were modest relative to the large interindividual variation (9.4-13.9dB IQRs). Individual ranges, averaged across sliders for i2 and i3, were correlated with trial duration (Spearman’s *ρ* = 0.48, 0.64 & 0.64 for i1, i2 and i3, respectively; all *p* < 0.01) . Like trial duration, the role of the individual in the range explored was supported by significant correlations between ranges using different sliders (Spearman’s *ρ* = 0. 75-0.96; all *p* < 0.001), as shown in the middle panels of Figure 5B. Unlike trial duration, there was no significant effect of repeat on range explored[χ^2^_(3,120)_= 5.29; *p* = 0.15]. Individual ranges were positively correlated with reliability across all sliders (Spearman’s *ρ* = 0.41; *p* ≪ 0.001); that is, variability in gain endpoints increased with the overall range in gains explored.

For each slider, there were significant differences between clusters in the ranges explored [χ^2^_(2,38)_= 8.72-13.85, all *p* < 0.05; ε^2^ = 0.21-0.34]. C3, which preferred relatively greater low-frequency gain, explored greater ranges than C2, which preferred gain closer to the initial reference (median Δ = 8.8-13.4dB; *z* = 2.64-3.25;all *p* < 0.05). C3 ranges were also greater than C1, which preferred relatively lower overall gain, but only for i2LF, i3LF and i3HF sliders (median Δ = 10.2-12.8dB; *z* = 2.94-3.45; all p < 0.05). There were no differences in ranges explored between C1 and C2 (*z* = 0.16-1.97;all *p* > 0.05).

##### Reversal rate

There were significant differences in reversal rate due to both interface [χ^2^_(2,80)_= 55.91;*p* ≪ 0.001; Kendall’s *W* = 0.35], with a greater rate for i1 than i2 or i3(*z* = 5.51 & 5.54, respectively; both *p* ≪ 0.001), and stimulus [χ^2^_(2,80)_ = 12.63; *p* = 0.0018; Kendall’s *W* = 0.08], with a lesser rate for speech than speech in noise and music (*z* = -2.97 & -3.53; both *p* < 0.01). While there was also a significant main effect of cluster on reversal rate for i2 [χ^2^_(2,38)_= 6.26; *p* = 0.044 ; ε^2^ = 0.15], none of the pairwise differences were statistically significant when corrected for multiple comparisons. As shown in the bottom row of Figure 5B, individual reversal rates were highly correlated across interfaces (*ρ* = 0.75-0.90; all *p* ≪ 0.001). Additionally, reversal rates were correlated with ranges explored for i2 and i3 (*ρ* = 0. 49-0.58; all *p* < 0.01). Unlike ranges, however, reversal rates were not correlated with self-adjustment reliability (*ρ* = -0.22–0.30; all *p* > 0.05).

#### Self-adjusted sound ratings and interface preferences

At the end of each trial, participants rated the adjusted sound while it played on a discrete scale of 1-5 stars. The slight majority of ratings, 59%, were 5 stars. We examined the effects of interface and stimulus type on ratings using Friedman’s test with Wilcoxon signed rank post-hoc comparisons. Median ratings for the three interfaces ranged from 4.2-4.5 stars and were not significantly different from one another [χ^2^_(2,80)_ = 5.54; *p* = 0.063]. There was, however, a significant main effect of stimulus [χ^2^_(2,80)_= 24.15;*p* < 0.001; Kendall’s *W* = 0.23], as speech in noise adjustments were rated lower than music or speech-in-quiet (*z* = -3.30 & -4.32, both *p* < 0.01). There was also a difference in ratings by cluster using Kruskal-Wallis ANOVA [χ^2^_(2,38)_ = 10.73; *p* = 0.0047 ; ε^2^ = 0.26], with post-hoc Wilcoxon rank-sum tests indicating C2 (endpoints relatively near starting point) had significantly higher ratings than C1 andC3 (*z* = 2.48 and 2.89, respectively; both *p* < 0.05).

After the self-adjustment task was completed, participants were asked which interface they preferred and to briefly explain why. One participant gave incongruent responses and was excluded from the analysis. The majority (80 %) of participants preferred either i2 or i3 (*n* = 14 & 18, respectively). The remaining participants equally chose either i1 or had no preference (both *n* = 4) . There was no relationship between cluster and interface preference. The open-ended responses were qualitatively analysed for themes related to interface preference. A prominent preference theme was effectiveness (*n* = 25 participants); when desiring “greater control” to decrease background noise, increase musical bass and/or improve sound quality, i1 was not preferred. When desiring ease or efficiency (combined *n* = 10), i3 was not preferred. For those who questioned the usefulness of the i3HF slider (e.g., “didn’t do anything… suspects it sometimes has no function”) or desired a “happy medium” (*n* = 10), the preferred interface was i2. Reasons for no preference included “doesn’t matter”, “doesn’t improve sound”, and “not sure; it’s good to have variety.”

### Sound-matching

#### Matching accuracy & reliability

Matching accuracy was measured as participants’ median absolute error (| Δ|) across four repetitions per slider and stimulus type, and matching reliability was measured as the standard deviation of the signed error σ(Δ) across the four repetitions. Results are shown in Figure 6. The effects of stimulus type and slider on sound-matching accuracy and reliability were assessed using Friedman’s test with Wilcoxon signed-rank post-hoc comparisons. There was no main effect of stimulus type on either sound-matching accuracy or reliability [χ2(2,62) = 1.69 & 1.94, respectively; both *p* >> 0.05], hence sound-matching results were pooled across stimulus types. (i.e., each individual matching accuracy and reliability were calculated across 12 trials, the four repeats of the three stimulus types). In terms of accuracy, there were significant differences in matching error between sliders [χ^2^_(5,155)_ = 112.45; *p* < 0.001; Kendall’s *W* = 0.70]. Errors were significantly smaller for i1 (median |Δ| = 1.7dB) than all other sliders (*z* = -3.347 - -4.937; all *p* < 0.05) except i2LF (*z* = -1.36; *p* > 0.05) . Errors were significantly smaller for sliders controlling low frequencies (i2LF & i3LF) than those controlling mid and high frequencies (i2HF, i3MF & i3HF; *z* = -3.011– -4.937; all *p* < 0.05). Moreover, errors were smaller for i2HF compared to i3MF and i3HF (z = -3.33 & -4.77; both *p* < 0.05) and for i3MF compared to i3HF (*z* = -4.43; *p* < 0.001). There were also significant differences in matching reliability between sliders [χ^2^_(5,155)_= 126.29; *p* < 0.001 ; Kendall’s *W* = 0.79]. The pattern was very similar to accuracy – reliability decreasing with increasing number of sliders and centre frequency – as individual matching accuracy and reliability were highly correlated (*ρ* = 0.94; *p* ≪ 0.001). For i3HF, median accuracy and median reliability were near and worse, respectively, than what performance would have been without adjustment from the ±5-12 dB baseline (8.5 & 8.7 dB for matching accuracy & reliability, respectively; red dotted lines in Figure 6) ; that is, participants would have achieved similar performance not adjusting the slider (n.b., they had to interact with all sliders to complete each trial).

**Figure 6.**
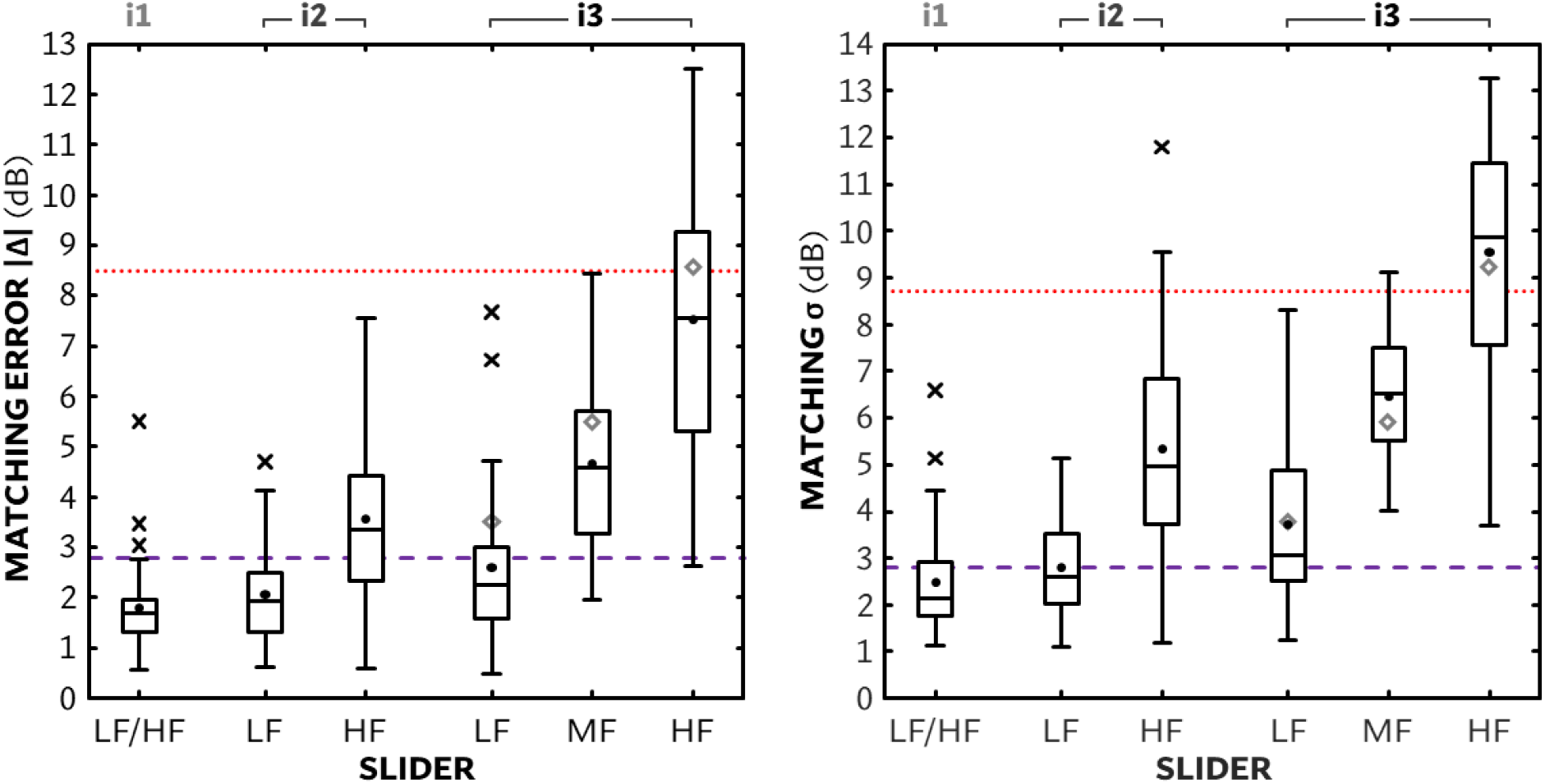
Left panel shows sound-matching error (absolute difference of adjusted-to-match gain and reference gain) for each interface/slider. Right panel shows sound-matching standard deviation (σ) for each interface/slider. Dots show means; dashed purple line shows median self-adjustment σ for comparison; red dotted lines show performance without adjustment from the ±5-12 dB baseline. Grey diamonds show median preference JNDs for 6-s stimuli in Whitmer et al. (2022).

#### Sound-matching behaviours

Sound-matching behavioural measures were calculated and analysed in the same manner as the self-adjustment behaviours described above. Like the self-adjustment task, sound-matching trial durations increased with number of sliders per interface [χ^2^_(2,62)_= 62.06; *p* < 0.001; Kendall’s *W* = 0.39] with medians of 22.7s for i1, 34.3 s for i2 and 53.3s for i3.Within each interface, there was no effect of stimulus on trial duration [χ^2^_(2,62)_ = 0.56–3.94; all *p* > 0.05]. Trial durations were only correlated with absolute error for i2 (*ρ* = -0.52; *p* = 0.0024). Individual ranges explored were correlated across sliders (*ρ* = 0.52–0.90; all p < 0.05). Ranges with each slider were negatively correlated with absolute errors (*ρ* = -0.49; *p* ≪ 0.001), but by slider, there was only a negative correlation between ranges and errors for the higher frequency sliders that also had relatively greater error: i2HF, i3MF and i3HF (*ρ* = -0.62 – -0.80; all *p* < 0.01). That is, greater ranges explored were associated with smaller errors (i.e., more accurate matching) when matching was more challenging.

### Relationship of sound-matching and self-adjustment

#### Sound matching accuracy and self-adjustment reliability

If matching performance reflects self-adjustment reliability, it may provide a useful indicator of an individual’s effective use of self-adjustment. Conceptually, the ability to match one sound to another would be related to more reliable adjustments to preference, hence we look here at the associations between individual matching accuracy and self-adjustment reliability. The left panel of Figure 7 shows self-adjustment endpoint standard deviations (reliability) as a function of sound - matching absolute error (accuracy); data points are color-coded to indicate cluster membership based on the self-adjustment endpoints. While there was a significant positive correlation between matching accuracy and self-adjustment reliability for i1 (*ρ* = 0.51; *p* = 0.016), there was no overall correlation (*ρ* = 0.13;*p* > 0.05) . By cluster, those who preferred more relative LF (C3) were generally more accurate but less reliable, although there were only six participants in this cluster with results for both tasks. There were otherwise no indications of any pattern in the results across the two tasks, indicating that the ability to match one sound to another is not a useful predictor of the ability to adjust to one’s preference.

**Figure 7.**
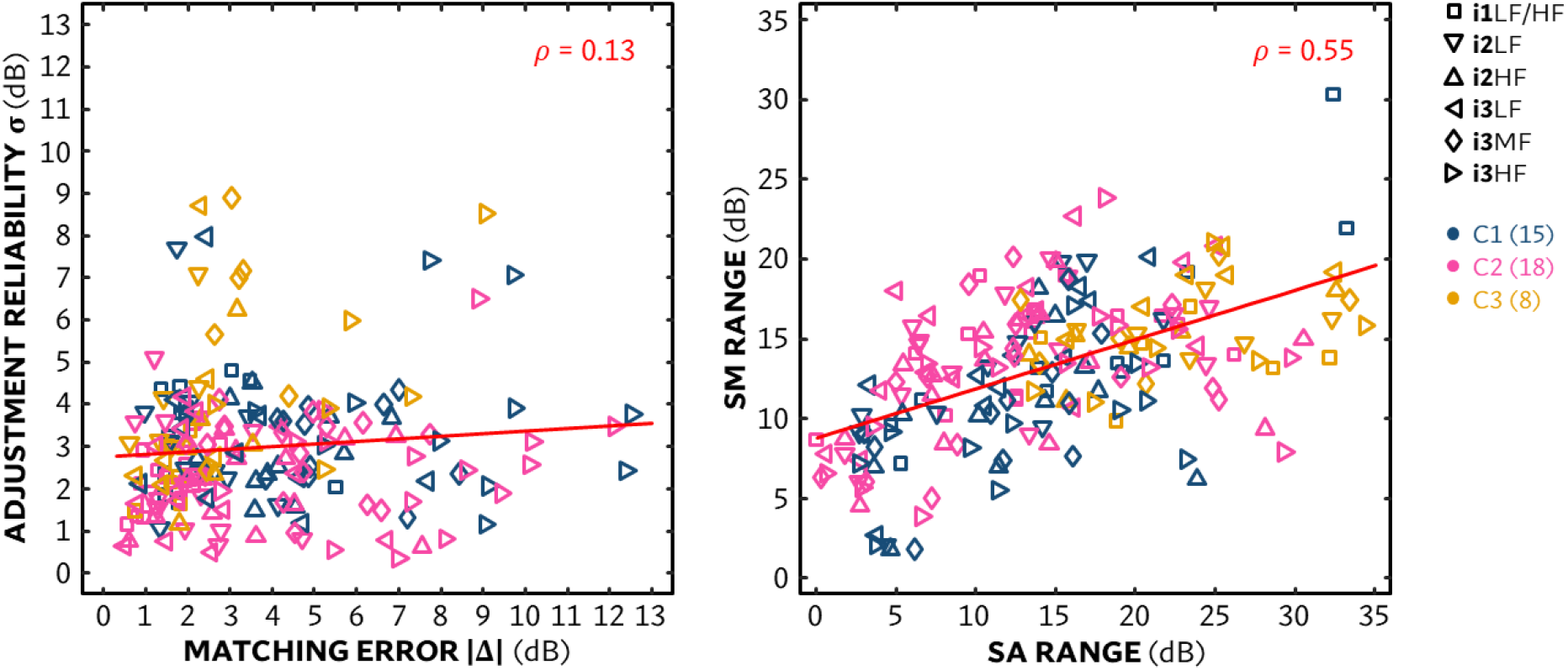
Left panel shows self-adjustment reliability as a function of sound-matching error for each individual with each interface/slider (symbol)Participant cluster is indicated by colour. The red line represents the insignificant linear regression of all data. Right panel shows sound-matching range as a function of self-adjustment range for each individual with each interface/slider (symbol).

#### Exploration of behavioural similarities and differences

The ranges explored per slider were relatively consistent within participants (middle panels of Figure 5B), and there were slider-specific correlations between the range explored per slider and how accurately individuals matched its gain to the reference sound. However, the overall range explored was smaller in the sound matching task compared to the self-adjustment task. This raises the question of whether interaction behaviour changes across tasks; specifically, to what extent the range explored in sound matching corresponds to that in self-adjustment. The right panel of Figure 7 shows the ranges explored across tasks. There was an overall significant positive correlation (*ρ* = 0.55; *p* ≪ 0.001), as well as positive correlations for all sliders (*ρ* = 0. 42–0.79, all *p* < 0.05), indicating that participants who explored wider ranges in self-adjustment tended to do so in matching as well. When observing the distribution of clusters across ranges explored, it is evident that the ranges from C3 have the tendency to be larger for both tasks, although the low number *n* =(6) of participants in this cluster that had results for the sound-matching task limit the ability to make inferences or draw conclusions.

## DISCUSSION

This study explored the effects of three self-adjustment interfaces differing in number of controls and three stimulus types on the self-adjustment of FGR, both in terms of participants’ preferred gains and the adjustment process. It also evaluated participants’ subjective preferences for an interface and whether a range of participant characteristics could explain aspects of individuals’ self-adjustments. However, only clustering individuals by self-adjustment endpoints yielded insights connecting some behaviours to adjustment endpoints. These clusters did not have clear associations with the measured participant characteristics in this sample. An analogous sound-matching task yielded PSEs for FGR changes but did not provide any insights into how or how well the same participants performed the self-adjustment task.

### Self-adjustment endpoints

Average self-adjusted gain endpoints broadly agreed with what has been observed in previous studies. Listeners with hearing loss generally chose less overall gain than their audiometric fittings over many generations of hearing aids and gain prescription formulae (e.g., Smeds et al., 2006 a/b; Dreschler et al., 2008; Mueller et al., 2008 ; Keidser & Alamudi, 2013; D’Onofiro et al., 2019) and relatively less gain (0.7–1.2 dB)in noisy listening conditions (i.e., the speech-in-noise stimulus; cf.Cox & Alexander, 1991; Keidser et al., 2005 ; Nelson et al., 2018 ; Kurşun et al., 2025a). In contrast, participants in Gößwein et al. (2023) preferred more gain in noisy listening conditions but also in conditions where the speaker characteristics changed. Average endpoints in the current study had more low-frequency relative to high-frequency gain than prescription, which also agreed with numerous prior studies (e.g., Kuk & Pape, 1992; Dreschler et al., 2008; Keidser et al., 2008; Nelson et al., 2018;Caswell-Midwinter & Whitmer, 2021; Vaisberg et al., 2021; Gößwein et al., 2023). Like several of those previous studies, however, the starting prescription (NAL-R) prescribes relatively little low-frequency gain compared to other prescriptions (e.g., Moore et al., 1999). The preference for greater relative low-frequency gain could then be considered compensation for an inadequate prescription, although the REIG at 250 and 500 Hz for the majority (78%) of participants’ hearing aids was below this same prescription.

Self-adjustments were relatively reliable within individuals insofar as the standard deviation across repeated adjustments (median 2.8 dB) was similar to studies comparing repeated adjustments (Elberling & Vejlby Hansen, 1999; Dreschler et al., 2008; Nelson et al., 2018; Mackersie et al., 2020), and relatively stable across interfaces and stimuli and repeated blocks (Kursun et al., 2025b). Based on findings of Nelson et al. (2018), it was hypothesised that speech in noise would produce more reliable self-adjustments than speech in quiet, but stimulus type did not have a statistically significant effect on self-adjustment reliability in the current study . Unique segments were used for each repetition of a stimulus, hence our measure of reliability, the endpoint standard deviation, could have been inflated by using a unique segment for each repetition .However, the segments were acoustically similar as contiguous music excerpts from the same piece, and neither reliability nor interface effects differed as a function of stimulus type. Nonetheless, there was modest spectral variation of approx. 1 dB across excerpts of each stimulus type that could have modestly inflated endpoint standard deviations. As i1 constrained adjustment (i.e., offered no overall level adjustment), it may be that its reliability was adversely affected by participants not being able to achieve a preferred FGR, but there was no significant difference in reliability between clusters that would have been expected to be differentially affected by i1’s constraints. Additionally, reliability was invariant to the frequency band, unlike sound-matching accuracy. This invariance could indicate that self-adjustment of high-frequency gain may be influenced by factors beyond acoustics alone, such as expectations or previously found solutions.

### Clustering of self-adjustment endpoints

Self-adjustment endpoints varied between individuals over a wide range spanning up to 29 dB (cf. ranges up to 40 dB in Nelson et al., 2018). This individual variation did not correspond to their own hearing aids’ REIGs at any corresponding frequency band. The large differences between how individuals adjusted puts into question the relevance of group averages for any individual, even though each participant began from an FGR based on their individual audiogram. To better characterise these individual differences, we clustered participants’ mean self-adjustment endpoints. Exploratory cluster analysis suggested three relatively distinct patterns: (1) less overall gain (*n* = 15, or 37% of participants), (2) close to the initial baseline (*n* = 18, 44%), and (3) more bass, less mid and/or treble (*n* = 8, 19%).

Clusters did not significantly differ in BE4FA, age, or answers to the any of the questionnaires (IOI-HA, TRI, BFI, LOC), similar to Perry et al. (2019), but did show differences in ranges explored. In particular, C3 explored a greater range than the other clusters. C3 preferred FGR was more complex than the other clusters, involving a relative difference between the three sliders. To find this relationship could have required larger adjustments to discover that relationship by assessing the unlabelled sliders’ functions. This connection between range explored and self-adjustment endpoints contrasts with Gößwein et al. (2024), who clustered participants based on the range they explored on a 2-D surface interface that controlled overall gain on the y-axis and spectral tilt on the x-axis, and those clusters were not linked to different self-adjustment endpoints. Range in that study, though, was based on surface area, hence to cover a large range (surface area), their participants would have been required to explore extremities of both parameters together (i.e., listening to both low- and high-frequency spectral tilts at both low and high overall gain), which may not have been a logical solution for that study.

Differences in these self-adjustment endpoint clusters could have been driven by differences in what individuals aimed to achieve or which criteria they applied during adjustment. Participants in Gößwein et al. (2023) adjusted to relatively greater overall gain when they were instructed to adjust the sound to maximise speech understanding relative to adjusting to maximise listening comfort. In Keidser et al. (2005), participants on average adjusted the spectral tilt for relatively greater bass gain of noisy stimuli when instructed to adjust for speech understanding, even when the SNR in the low frequency bands was negative, and increased bass gain would not have provided an objective improvement (i.e., in traffic noise; cf. Skinner & Miller 1983). Boothroyd et al.(2022) found that when participants were not instructed to adjust to any one criterion, they reported at the end of the adjustment task to use a variety of criteria. In the current study, participants were also not instructed to adjust to any criterion, and as described in the Rating of self-adjusted FGRs section below, when asked after adjustment what was their preferred interface and why, numerous reasons were given.

### Self-adjustment behaviours

Self-adjustment durations increased with increasing number of sliders per interface, each slider adding roughly 4 s to the i1 median trial duration of 16.5 s. These durations are relatively fast compared to those previously observed in the literature, averaging approx. 45-60 s (Dreschler et al., 2008; Rennies et al., 2016; Kliesch et al., 2023). While repeated adjustments have previously been generally faster than initial adjustments (Boothroyd & Mackersie, 2017; Boothroyd et al., 2025; Gößwein et al., 2023; Mackersie et al., 2020), the effect here was small (2 s faster; cf. 3 s faster in Dreschler et al. 2008), although this may be due to having six practice trials. Trial duration varied with stimulus type; adjusting speech in noise took longer than music or speech. The tendency for longer adjustment durations in noisy conditions was also reported by Nelson et al. (2018), Perry et al. (2019) and Walravens et al. (2020), although no differences in time spent adjusting speech in noise vs. in quiet was found by Mackersie et al. (2020), possibly due to a differece in methods, such as allowing the initial response to be accepted without adjustment. Additionally, ranges explored and reversal rates for speech in noise were also greater than those for speech in quiet, although not significantly different than the music stimuli. What was apparent through the subjective preferences given at the end of the task is that in noisy situations, some participants were attempting to use FGR adjustments to reduce the noise. The longer trial duration for speech in noise could be due to the difficulty in achieving this aim given the limited number of broad filters and common spectral overlap between the speech and noises used in the current and past studies. That is, the ease and effectiveness of the self-adjustment is dependent on the aims, partially determined by the stimuuls. These results counter the previous hypothesis that a more spectrotemporally rich signal such as speech in noise would be easier to adjust to preference based on its better (lower) FGR JND (Caswell-Midwinter & Whitmer, 2019a).

As shown in Figure 5B, individual trial durations, ranges explored and reversal rates were highly correlated across interfaces and sliders, indicating that an individual will approach different interfaces with the same behaviour. Unlike Gößwein et al. (2024) who found no relationship between exploration on a 2-dimensional gain-frequency grid and FGR endpoints, ranges explored in the current study were indicative of preferred FGR cluster, insofar as those preferring greater relative bass (C3) generally had greater ranges. There were, though, also a few participants who explored, at least with some interfaces, a relatively wide range but had preferences near the starting reference (C2), indicating that the wide range could have been indicative of exploring the extremes– bracketing – as an assurance that the starting reference was near their preference . It is also possible that some differences in individual ranges are due to differences in uncertainty. When participants are unsure on their preference or cannot clearly perceive acoustic changes during self-adjustment, they may revert to the starting point as they consider it a neutral or non-detrimental choice (Tversky & Kahneman, 1974). Future research should look at how the range is affected by different(momentary) aims in self-adjustment to foster greater interpretation of self-adjustment gestures.

### Rating of self-adjusted FGRs

Participants evaluated the sound of their self-adjusted FGRs by rating the adjusted sound as it played on a discrete 1-5 scale directly after the adjustment. These ratings after adjustment exhibited strong ceiling effects, and there was no effect of interface one possible explanation is that immediate evaluation was biased by psychological ownership of the adjustment (e.g. Pierce et al,. 2003; Convery et al., 2011). Such bias has been suggested to account for greater satisfaction with the sound of a s-elf adjusted hearing device compared to one set by audiology best practice (Sabin et al., 2020). Unlike Sabin et al. (2020), who did not find an effect of listening environment, there was an effect of stimulus type in the current study on ratings. Adjustments of speech-in-noise stimuli were rated lower than those of the two other stimulus types, which agrees with other research showing that hearing-aid sound quality is generally perceived as worse in noisy conditions (e.g., Arehart et al., 2010; Neher, 2014; Lundberg et al., 2020). Additionally, previous studies using speech-in-noise stimuli found no improvement by self-adjustment over prescriptive settings (Nelson et al., 2018; D’Onofrio et al., 2019; Gößwein et al., 2023); as noted in the previous section, FGR adjustments are unlikely to improve speech perception, which in turn can influence the sound quality. Such results suggest that adjusting other settings outwith FGR (e.g., noise reduction) within a user interface may be necessary for personalisation when listening to speech in background noise.

At the end of the self-adjustment task, participants also chose their preferred interface of the three used and gave reasons for their preference. Unlike Dreschler et al. (2008) where participants gave ratings for each interface, which yielded very similar results across interfaces for most questions, participants in the current study provided reasons for their preference. Ease and efficiency were associated with i1 and i2, whereas effectiveness, manifest in any desire to control the sound generally or stimulus-specific aspects of it (e.g., bass in the music, noise in the speech in noise), was associated with i2 and i3. Additionally, when no perceptual change was reported with the i3 HF slider, i2 was preferred. Based on these reports as well as the preferred endpoints, i2 can be considered a “happy medium” that satisfies the ease, effectiveness and efficiency needed to make a an FGR self-adjustment method useful, but the most preferred interface was i3. Additionally, participants mentioned, albeit rarely, a desire for different interfaces in different situations. Even with limited scene-awareness, a method could tailor the interface to the relevant adjustments for that scene, although this would need to be leveraged against the potential cost in ease of using different scene-determined controls. These results indicate how individuals apply different criteria, and that criteria are weighted not only individually but ephemerally, depending on the initial immediate impression of the sound (e.g., speech understanding may not be the most important criteria if speech can be understood well without any adjustment from baseline). These myriad possible aims and their dynamic relationship within the self-adjustment process warrant a more qualitative approach (e.g., cognitive interviews on self-adjustments) to understand adjustment criteria.

### Relation of self-adjustment and sound matching

One of the aims of the current study was to see if sound matching can be an indicator of the effectiveness of self-adjustment, as inferred from the reliability of repeated self-adjustments. There was, though, a lack of correlations between matching accuracy and self-adjustment reliability, as well as a frequency dependence in sound-matching, with higher frequency band matching being more difficult than lower-frequency matching (cf. Caswell-Midwinter & Whitmer, 2019b), that was not evident in self-adjustment. There were positive correlations between the two tasks for both the trial durations and ranges explored (right panel of Figure 7), indicating that an individual applies a similar behaviour to both tasks, but these behaviours were correlated with performance on the tasks in opposing ways. Increased range and duration in self-adjustment was correlated with increased variation in endpoints (poorer reliability), whereas increased range in sound matching was correlated with decreased error and variation. These differences suggest a perceptual distinction between the two method-of-adjustment tasks despite using the same interfaces, stimuli and having acoustically equivalent targets (i.e., setting the sound-matching targets as the self-adjustment endpoints). Participants who were highly accurate in sound matching but showed lower reliability in self - adjustments demonstrated an ability to perceive differences and operate the interfaces meaningfully during the matching task. These participants may be just as capable of self-adjustment, but either may simply have broader FGR tolerances for preference (as hypothesised by Dreschler et al., 2008) or changed their preference criteria (i.e., found multiple FGR solutions) across self-adjustment trials. This preference dynamism may explain how the adjustment reliability for sound matching (with an external reference) differed from the same participants’ self-adjustments (to their internal reference) even though the endpoints of adjustment were identical. Hence, sound matching may not be a useful indicator for self-adjustment proficiency. For self-adjustment, the positive correlation between endpoint standard deviation and range explored potentially indicates that adjusting over a larger range is a sign of uncertainty, either in preference or in the functionality of the adjustment. The thought processes behind exploration in self-adjustment warrant further study.

### Limitations

The current study has been conducted in the lab using headphones and recorded speech and music stimuli presented in a loop. These conditions differ substantially from dynamic and interactive listening situations in the real-world but also differ acoustically when listening through hearing aids. The adjustments applied in this study involved substantial linear gain changes (±18 dB) across an extended frequency range, which may be outside what can be achieved in hearing aids while maintaining stable gain. Previous studies with a substantial adjustable range have also produced large interindividual variation in the lab (Nelson et al., 2018; Perry et al., 2019), whereas real-world adjustments with the same interface showed less interindividual variation as well as less deviation from prescription (Sabin et al., 2020). Self-adjustments made in the lab have been found to be only a fair predictor of real-world adjustments (Punch et al., 1994). Possible reasons for this arise from the more dynamic and varied rea-l world listening environments and situations, not only acoustically but in terms of their dynamic and interactive nature (i.e., conversation; e.g., Brungart et al., 2014; Beechey, 2022). The adjustable range available to hearing-aid users outside of the lab needs to be sufficient to provide meaningful differences, not only as preferred gain endpoints but also for exploration of those endpoints, as shown by the ranges explored in the current study. An adjustable range of at least ±8 dB has been recommended by Cox and Alexander (1991), and comparable ranges have been used in past research (Hornsby & Mueller, 2008; Mueller et al,. 2008). A smaller range, such as the ±6 dB in some current hearing-aid smartphone apps, may not allow bracketing. Given that preferred settings found in the current as well as past studies being a reduction in gain from prescription, in particular high - frequency gain, future self-adjustment tools could limit the incremental range in those frequencies but extend the decremental range without the issue of gain instability (i.e., feedback).

The current study also used stimuli with greater bandwidth, including lower frequencies (< 250 Hz), than those typically available in hearing aids, especially open-fit devices commonly chosen for individuals with mild to moderate losses. Furthermore, hearing aids include other signal processing parameters, most notably wide dynamic range compression, directionality, feedback suppression and noise reduction, some of which may interact with user preferences in complex ways in that they may not converge or prdouce reliable preferential parameter settings (Franck et al., 2007). The current study purposively used an antiquated linear prescription formula, NAL-R, as a starting point to promote self-adjustment, but does not reflect current hearing-aid gain prescriptions. The choice of NAL-R may have led to exaggerated adjustments away from baseline, although Nelson et al. (2018) also reported substantial and varied adjustments away from the more current NAL-NL2. What was perhaps more surprising was how many participants (clusters 1 & 2) preferred an FGR with a similar spectral shape to NAL-R. That is, whereas the results across participants (Figure 3 left panel) showed a relative bass emphasis similar to previous studies, the clusters here show that this emphasis was driven by a subgroup, and that the majority (C1 & C2) only deviated approx. +2 dB on average in LF from the higher-frequency emphasis of the NAL-R prescription. Regardless, future studies should strive to include more of the processing and modern prescriptions that reflect rea-l world hearing-aid fittings. Adjustments in the current study required interaction with all sliders, and adjustments were repeated within a session, which is likely different from how individuals would use and interact with self-adjustment in real-world situations. The lack of relationship between participants’ current hearing-aid gains, their hearing-aid experience and their self-adjusted gains in the current study could be related to these methodological differences between the current study and clinical fitting . These differences between self-adjusted and REIGs could also be construed as an appeal to promote self-adjustment in the clinic, but further research is necessary to move beyond an absence of evidence.

The study sample size as a whole (*n* = 41) was reasonably powered relative to previous studies, but the post-hoc clusters had small participant numbers, particularly C3. What is crucial to note is that the clusters only differed in their preferred FGRs, and not on any other individual metric we applied (i.e., age, hearing loss, characteristics potentially related to self-adjustment). The initial impression of the sound, as well as the criteria an individual applies, may play an important role in how adjustment endpoints are determined, and future studies need to be able to capture these impressions in some capacity. Quantitatively, such endeavours will need to consider how different criteria are weighted by each individual and ensure that stimuli are personally relevant.

## CONCLUSIONS

Generally, the group-level patterns in self-adjusted FGRs in the current study were broadly consistent with previous findings in the literature, with relative more bass than mid or treble and less overall gain when possible relative to an individualised NAL-R prescription. Also, like previous studies, there was substantial interindividual variation in self-adjustment endpoints. To characterise this variation, an exploratory cluster analysis was performed on participants’ self-adjustment endpoints. The analysis suggested three basic gain preferences relative to an individual prescription : less gains across frequencies (C1), gains similar to the prescription (C2) and relatively more bass than mid/treble (C3) . The three clusters did not differ significantly in BE4FA, age or participant characteristics assessed with questionnaires potentially related to the self-adjustment of gain (IOI-HA, TRI, BFI, LOC). Low-frequency gain preferences distinguished the clusters, and the minimum average gain differences between clusters were approx. 6 dB, which is the maximum adjustable gain in some current hearing-aid apps. In terms of application, these clusters could be translated into three buttons that allowed greater changes than ±6 dB, it is also important to consider that these FGRs were derived through a method of adjustment that allowed participants to explore each control, and that different solutions may result through a limited selection of FGRs. If the aim of a self-adjustment tool is to produce a single set of FGR parameters, then it may be advisable to explore individuals’ range of acceptable settings and provide them with settings corresponding to the centre of that range.

In the context of the limited stimuli and fixed initial presentation level of the current study, the choice of method in self-adjustment can be seen as a series of trade-offs between the interface’s ease, effectiveness and efficiency. We found that reliability, a common measure of self-adjustment effectiveness, was not significantly different across the interfaces. A single-slider interface offered modest increases in efficiency and self-reported ease, as well as greater accuracy in sound matching, but was reported to be less effective at achieving preferred gains, especially for those wanting overall level control (i.e., C1), which was expected as i1 prevented overall gain adjustment. Increasing the number of sliders from two to three can be interpreted as a modest decrease in efficiency (i.e., a 4-s average increase in adjustment duration), but for effectiveness and ease, the results are mixed. Having a separate MF control in i3 offered increased effectiveness insofar as greater specificity in preferred gains for those who wanted more bass (i.e. C3) but for some participants, the HF slider in i3, controlling gain above 2.6 kHz, was reported to not produce any perceptible changes. Although an interface with three sliders controlling bass, mid and treble separately was the most preferred by participants on reflection, three sliders may only be preferable in contexts where users can perceive the effect of and effectively use the additional control, and ease and efficiency are less important. Otherwise, two sliders may be the better practical compromise. Given these trade-offs, future hearing-aid apps should offer flexibility, potentially allowing users to, in effect, personalise the method of personalisation with context-aware interface choices – or the linking of sliders (e.g., MF & HF) within the same interface– adaptable to factors such as user experience and listening environment.

Self-adjustments were generally reliable (σ = 2.8 dB). There was a small (0.4 dB) significant difference in reliability between the high-frequency sliders in the two - and three-slider interfaces, otherwise reliability was broadly consistent across interfaces and stimuli . Conversely, sound- matching performance decreased with increasing frequency of the filter and a greater number of controls per interface, although was also invariant to stimulus type. While it was initially assumed that sound-matching is the psychophysical equivalent of self-adjustment, with exception of i1, this study found no monotonic relationship between matching accuracy and self-adjustment reliability, suggesting that these tasks are psychophysically distinct. Consequently, sound matching cannot predict how reliably individuals would arrive at the same self-adjusted FGR. In particular, the discrepancy between reliable self-adjustments and inaccurate matches when adjusting high - frequency controls suggests that self-adjustment may be guided by other factors than acoustics alone. Additionally, some participants were relatively more accurate at matching yet were relatively less reliable in self-adjustments; reliability may not be the most appropriate measure of an individual’s ability to perform self-adjustment. If individuals have a broader range for equally preferrable FGRs, they may benefit from guidance on how to employ self-adjustment strategies such as bracketing that could help them to adjust the FGR to within this range. Such guidance requires additional qualitative research on the goals and aims hearing-aid users have when interacting with self-adjustment tools in order to promote easy, effective and efficient FGR personalisation.

## Data Availability

Anonymised study data is currently being prepared for repository submission

## ACKNOWLEDGEMENTS

This work was supported by funding from the Medical Research Council [grant number MR/X003620/1]; and GN Hearing A/S. Thanks to Prof. Pamela Souza and two anonymous reviewers for their constructive comments on a previous version of this manuscript . Thanks also to Prof. Graham Naylor, Gregory Olsen and Dr. Qi Yang for helpful comments throughout this work, Andrew Lavens and David McShefferty for technical assistance, and to all the participants.

